# Risk factors for severe COVID-19 among HIV-infected and-uninfected individuals in South Africa, April 2020- March 2022 – data from sentinel surveillance

**DOI:** 10.1101/2022.07.20.22277839

**Authors:** Sibongile Walaza, Stefano Tempia, Anne von Gottberg, Nicole Wolter, Jinal N. Bhiman, Amelia Buys, Daniel Amoako, Fahima Moosa, Mignon du Plessis, Jocelyn Moyes, Meredith L. McMorrow, Halima Dawood, Ebrahim Variava, Gary Reubenson, Jeremy Nel, Heather J Zar, Mvuyo Makhasi, Susan Meiring, Vanessa Quan, Cheryl Cohen

## Abstract

**Background:** Data on risk factors for COVID-19-associated hospitalisation and mortality in high HIV prevalence settings are limited.

**Methods:** Using existing syndromic surveillance programs for influenza-like-illness and severe respiratory illness at sentinel sites in South Africa, we identified factors associated with COVID-19 hospitalisation and mortality.

**Results:** From April 2020 through March 2022, SARS-CoV-2 was detected in 24.0% (660/2746) of outpatient and 32.5% (2282/7025) of inpatient cases. Factors associated with COVID-19-associated hospitalisation included: older age (25-44 [adjusted odds ratio (aOR) 1.8, 95% confidence interval (CI) 1.1-2.9], 45-64 [aOR 6.8, 95%CI 4.2-11.0] and ≥65 years [aOR 26.6, 95%CI 14.4-49.1] vs 15-24 years); black race (aOR 3.3, 95%CI 2.2-5.0); obesity (aOR 2.3, 95%CI 1.4-3.9); asthma (aOR 3.5, 95%CI 1.4-8.9); diabetes mellitus (aOR 5.3, 95%CI 3.1-9.3); HIV with CD4 ≥200/mm^3^ (aOR 1.5, 95%CI 1.1-2.2) and CD4<200/mm^3^ (aOR 10.5, 95%CI 5.1-21.6) or tuberculosis (aOR 12.8, 95%CI 2.8-58.5). Infection with Beta (aOR 0.5, 95%CI 0.3-0.7) vs Delta variant and being fully vaccinated (aOR 0.1, 95%CI 0.1-0.3) were less associated with COVID-19 hospitalisation.

In-hospital mortality was increased in older age (45-64 years [aOR 2.2, 95%CI 1.6-3.2] and ≥65 years [aOR 4.0, 95%CI 2.8-5.8] vs 25-44 years) and male sex (aOR1.3, 95%CI 1.0-1.6) and was lower in Omicron -infected (aOR 0.3, 95%CI 0.2-0.6) vs Delta-infected individuals.

**Conclusion:** Active syndromic surveillance encompassing clinical, laboratory and genomic data identified setting-specific risk factors associated with COVID-19 severity that will inform prioritization of COVID-19 vaccine distribution. Elderly, people with tuberculosis or people living with HIV, especially severely immunosuppressed should be prioritised for vaccination.

**Summary of article’s viewpoint:** Compared to the Delta variant, the Omicron variant was associated with reduced risk of mortality and Beta associated with decreased risk of hospitalisation. Active syndromic surveillance combining clinical, laboratory and genomic data can be used to describe the epidemic timing, epidemiological characteristics of cases, early detection of variants of concern and how these impact disease severity and outcomes; and presents a viable surveillance approach in settings where national surveillance is not possible.

## Background

Since its emergence in December 2019, coronavirus disease 2019 (COVID-19), caused by severe acute respiratory syndrome coronavirus 2 (SARS-CoV-2), has spread globally causing severe morbidity and mortality[1]. South Africa reported its first case of laboratory-confirmed COVID-19 on 05 March 2020, and experienced four epidemic waves during the study period[2]. By the end of November 2021, a high proportion of the South African population had some level of SARS-CoV-2 immunity. Seroprevalence in the catchment area of two surveillance sites ranged from 60%-70%, however it was estimated that <10% of cases were clinically diagnosed [3]. Observational studies, mostly conducted in high-income countries have described risk factors associated with increased risk of severe COVID-19 (hospitalisation and death), which included, older age, male sex and presence of comorbidities[4-6]. However, data on risk factors for COVID-19-associated hospitalisation and mortality in settings with high HIV or tuberculosis prevalence are limited. In South Africa in 2021, HIV prevalence was 19.5% among individuals aged 15-49 years, while tuberculosis prevalence was amongst the highest globally[7].

As SARS-CoV-2 evolves and new variants and lineages emerge, monitoring the impact they have on disease severity remains important. Sentinel surveillance programmes established to monitor influenza and other respiratory pathogens represent a potentially important platform for monitoring SARS-CoV-2 in low and middle-income countries, like South Africa, where resources for testing may be limited. South Africa’s vaccine program began on 17 February 2021, and as of 7 April 2022, 44% of individuals aged ≥18 years were fully vaccinated against SARS-CoV-2 (one dose of Johnson & Johnson or two doses of Pfizer-BioNTech)[8].

Using a well-established syndromic surveillance programme for influenza-like-illness (ILI)[9] and severe respiratory illness (SRI)[10-12], we aimed to describe clinical and epidemiological characteristics of persons with laboratory-confirmed COVID-19 and identify factors associated with COVID-19 hospitalisation or mortality.

## Methods

### Surveillance programmes

We enhanced our existing syndromic surveillance programme for influenza, respiratory syncytial virus (RSV) and *Bordetella pertussis* to include molecular testing and genomic surveillance for SARS-CoV-2 from 01 April 2020. We conducted active, prospective, clinic-based surveillance for ILI at 4 public health clinics in 3 provinces. In addition, we conducted active prospective surveillance for SRI at 8 public sector hospitals in 5 provinces; 4 hospitals were in the same catchment areas as the ILI clinic sites.

### Case definitions for enrolment

Individuals aged ≥ 15 years presenting to outpatient surveillance clinics meeting the ILI or suspected COVID-19 surveillance case definitions including any of the following:

i. ILI - fever (≥38°C and/or self-reported fever) AND cough AND symptoms ≤10 days
ii. Suspected COVID-19 - acute (≤14 days) respiratory tract illness OR other clinical illness compatible with COVID-19 (cough, sore throat, shortness of breath, anosmia or dysgeusia) OR physician-diagnosed suspected COVID-19 Individuals aged ≥15 years hospitalised at surveillance hospitals meeting the SRI case definition including any of the following:
iii. Individuals with physician-diagnosed lower respiratory tract illness (LRTI) or physician-diagnosed suspected COVID-19

Henceforth, all outpatients meeting the case definitions used at ILI sites will be referred to as ILI cases, and all hospitalised patients meeting any of the case definitions used at SRI sites will be referred to as SRI cases.

### Study procedures

The procedures for surveillance have been described previously[12-15]. Briefly, all patients presenting at ILI sites from Monday to Friday (08H00-16H00) or admitted at SRI sites from 17H00 on Sunday through 13H00 on Friday and meeting surveillance case definitions were eligible for enrolment. In addition, cases admitted at SRI sites over the weekend and testing positive for SARS-CoV-2 from testing conducted as part of clinical care were also enrolled if they met surveillance case definitions. Dedicated surveillance staff (surveillance officers (nurses) and research assistants) screened all outpatients or medical admissions, sought consent, and completed enrolment procedures if eligibility criteria were met. Enrolled hospitalised patients were followed until outcome (discharge, transfer or death). Case record forms were completed through structured interviews and by reviewing hospital medical records.

### Laboratory procedures

Nasopharyngeal swabs (Copan Italia, Brescia, Italy) were collected from study participants on enrolment, placed in universal transport medium, and stored at 4-8°C for transport on ice in a cooler box for testing at the National Institute for Communicable Disease (NICD) within 72 hours of collection. Nucleic acids were extracted from 200µl of transport medium using a MagNA Pure 96 automated extractor and MP96 DNA and Viral NA Small Volume v2.0 extraction kit (Roche Diagnostics, Mannheim, Germany). Extracts were tested for influenza viruses, RSV, and SARS-CoV-2 using real-time reverse transcription polymerase chain reaction (rRT-PCR). In addition, SARS-CoV-2 positive samples were tested by variant PCR and/or sequenced at NICD to ascertain their lineage/clade[16]. Additional information on rRT-PCR testing and sequencing methods are included in <u>supplementary material</u>.

HIV testing was ordered by attending clinicians as part of the standard of care. Pre-test counselling and bedside HIV testing was performed by surveillance staff for consenting patients not tested by the attending clinician. Among people living with HIV (PLHIV), no or mild HIV immune compromise was defined as CD4+ T-lymphocytes ≥200/mm^3^ and severe immune compromise as CD4+ T-lymphocytes <200/mm^3^.

### Statistical analysis

To assess factors associated with SARS-CoV-2-hospitalisation, only sites in provinces with ILI and SRI surveillance in the same catchment area were included in the analysis. All eight hospitals were included in the analysis of factors associated with SARS-CoV-2 infection and mortality. Logistic regression was used to assess factors associated with (i) SARS-CoV-2 infection among ILI or SRI cases at all surveillance sites; (ii) COVID-19-hospitalisation by comparing the characteristics of SARS-CoV-2-positive SRI cases (cases) to those of SARS-CoV-2-positive ILI cases (comparison group); and (iii) in-hospital mortality among SARS-CoV-2-positive patients with SRI.

The wave variable, defined as national weekly case incidence of ≥30 cases/100 000 (in-wave) and incidence <30 cases /100 000 (out of wave) and period variable, defined as time in months since the first documented case in South Africa to time of presentation, were included a priori in all models, to account for bias due to possible changes in enrolment of cases and clinical management during periods of increased transmission and changes in severity as population immunity increases as a result of infection or vaccination over time and improved treatment becomes available. Vaccination status was also included a priori as an important confounder on infection and severity.

To account for clustering by site, random effects were applied in all multivariable logistic regression models. Significant variables p<0.10 on univariate analysis were evaluated for inclusion in the multivariable models. Non-significant variables at p≥0.05 were dropped using manual backward elimination. Pairwise interactions were assessed by inclusion of product terms for all variables remaining in the final multivariable additive model. Stata version 14 (StataCorp Limited, College Station, TX) was used for analysis.

### Ethical approval

The ILI and SRI protocols were approved by the University of Witwatersrand Human Research Ethics Committee (HREC), reference M180832 and M140824, respectively. Additional approvals were received from other HRECs <u>supplementary material</u>. This activity was reviewed by CDC and was conducted consistent with applicable US federal law and CDC policy (see e.g., 45 C.F.R. part 46, 21 C.F.R. part 56; 42 U.S.C. 241(d); 5 U.S.C. 552a; 44 U.S.C. 3501 et seq).

## Results

From 01 April 2020 through 31 March 2022, 10 040 patients aged ≥15 years were enrolled from all sentinel sites, of whom 9781 (97.4%) had SARS-CoV-2 results. Of the cases with results, 2757 (28.2%) had ILI and 7025 (71.8%) had SRI. Among ILI cases, 10 (0.4%) were referred to hospital after consultation at an ILI site, and were excluded from further analysis. A total of 9771 patients were included in the analysis for description of factors associated with testing positive for SARS-CoV-2 (Table 1).

**Table 1:**
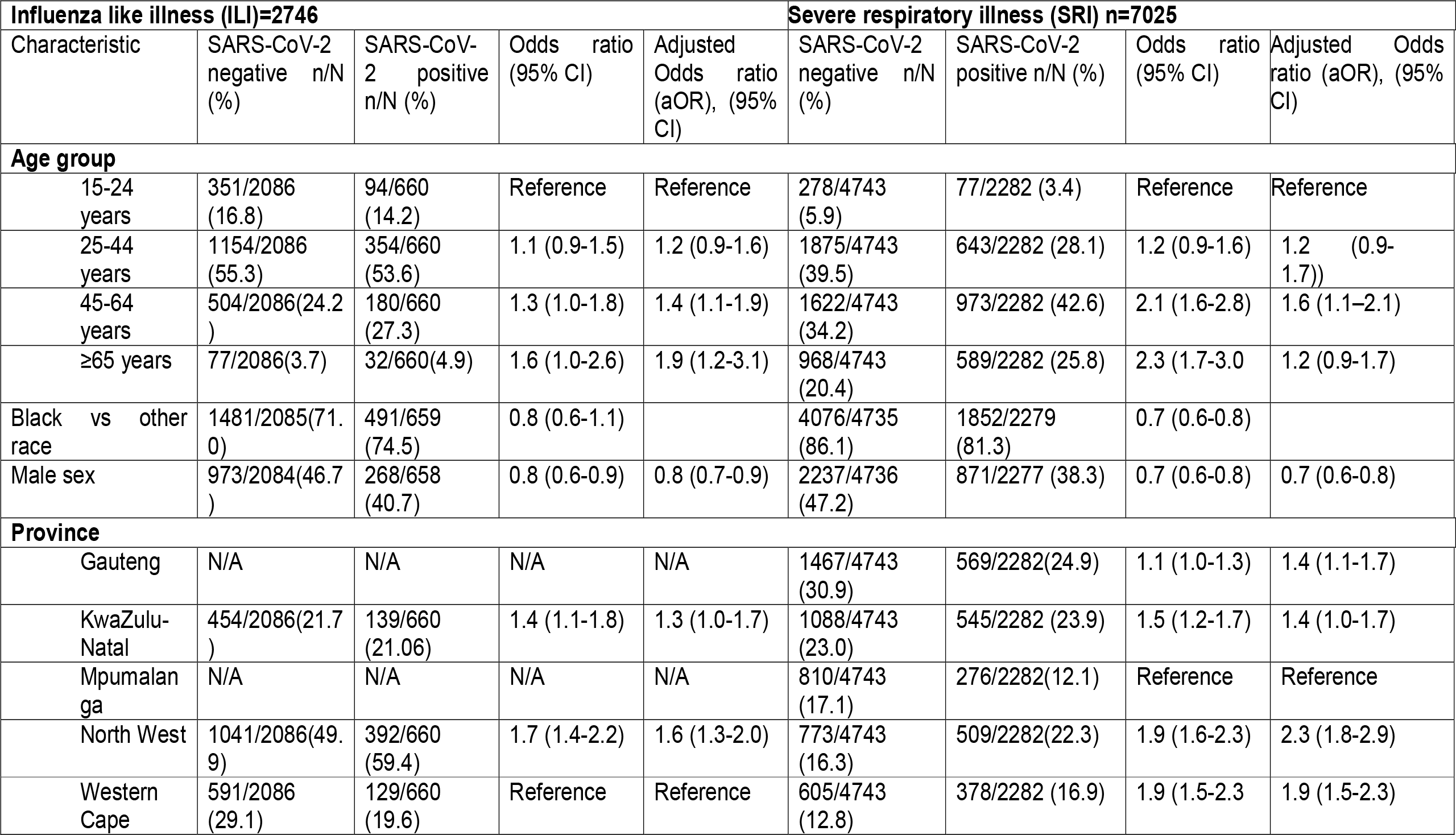
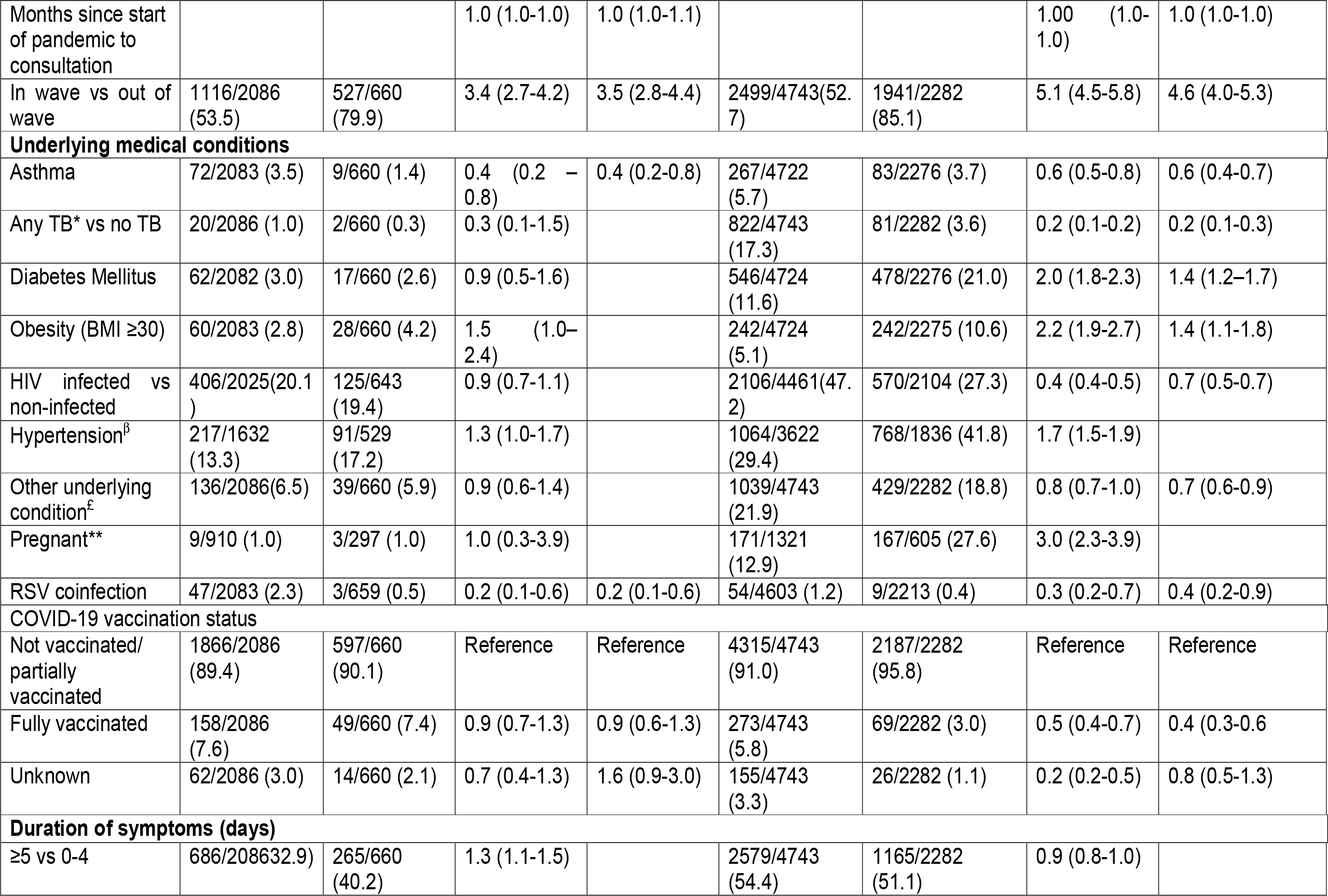
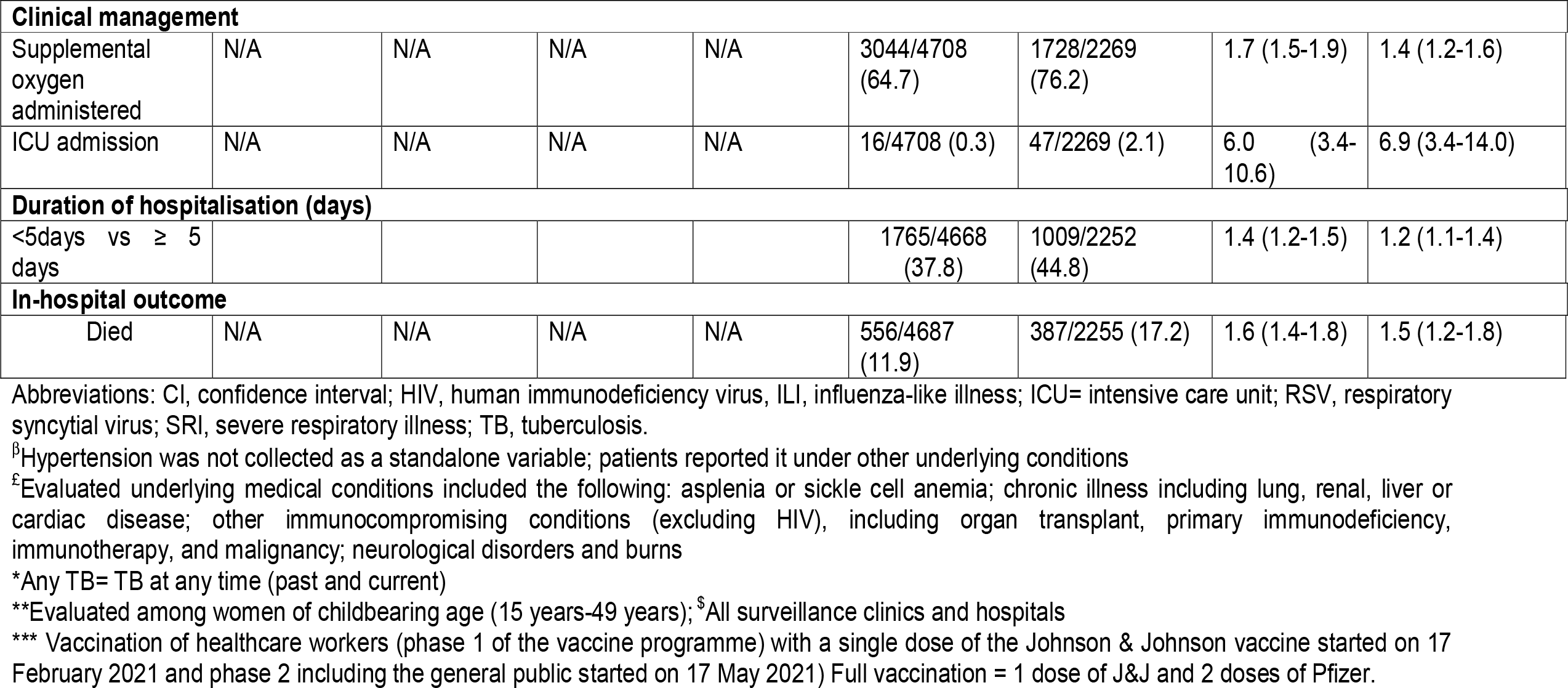
Demographic and clinical characteristics of patients with and without laboratory-confirmed (rRT-PCR) SARS-CoV-2 presenting with influenza like illness (ILI) or hospitalised with severe respiratory illness (SRI) at sentinel sites$ in South Africa, April 2020-March 2022 (N=9141)

SARS-CoV-2 was detected in 24.0% (660/2746) of ILI and 32.5% (2282/7025) of SRI cases. During the study period the COVID-19 epidemic in South Africa had four waves; predominated by SARS-CoV-2 Wuhan-Hu, Beta, Delta and Omicron (BA.1 and BA.2) variants, respectively (Figure 1a and 1b).

**Figure 1a:**
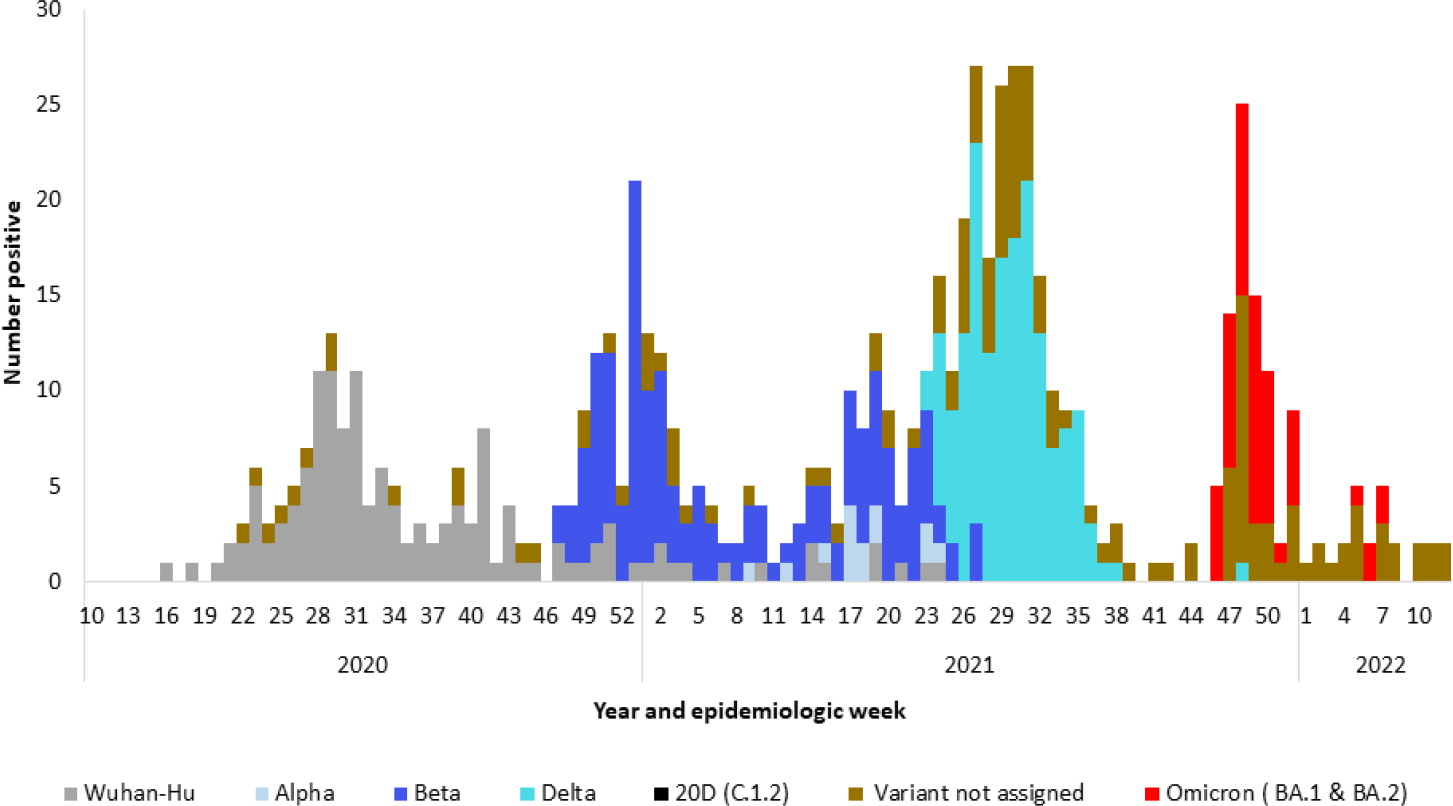
Number of laboratory-confirmed (rRT-PCR) COVID-19 cases aged ≥15 years by SARS-CoV-2 variant type and epidemiologic week, influenza-like illness surveillance, South Africa, April 2020-March 2022 Variant not assigned: no lineage assigned due to poor - sequence quality **OR** low viral load (ct≥35) **OR** variant PCR could not assign variant and no sequencing result

**Figure 1b:**
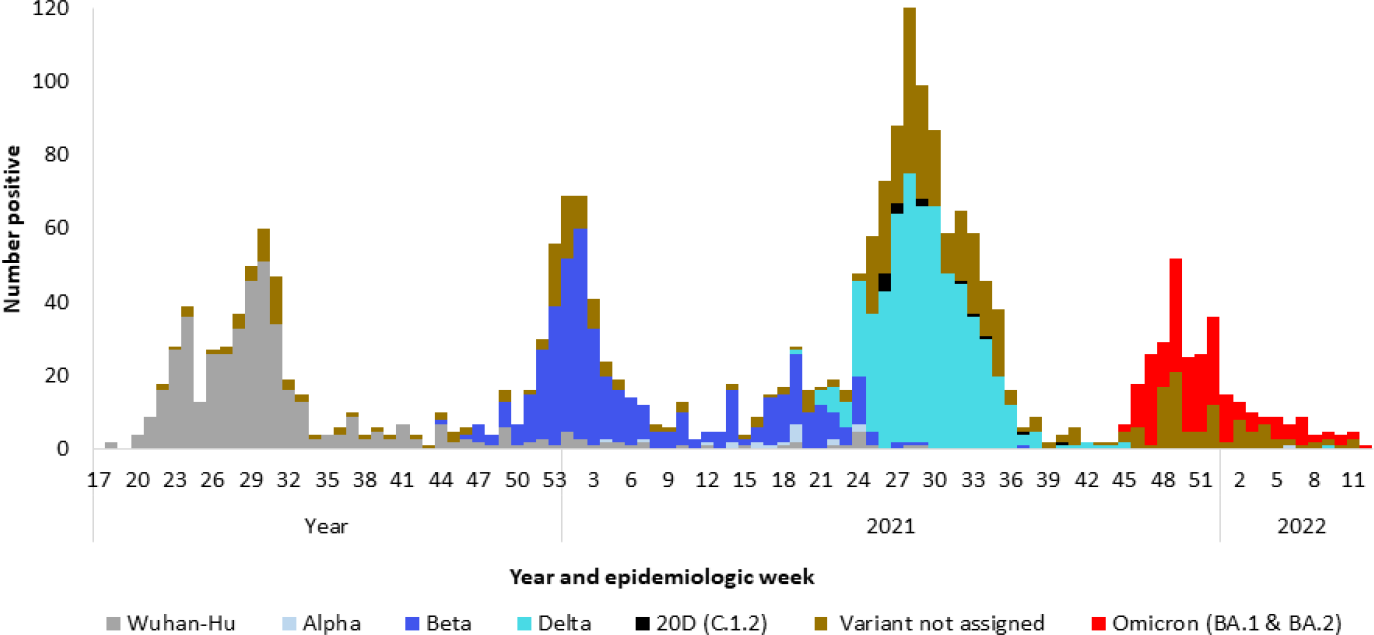
Number of laboratory-confirmed (rRT-PCR) COVID-19 cases aged ≥15 years by SARS-CoV-2 variant type and epidemiologic week, severe respiratory illness surveillance, South Africa, April 2020-March 2022 Variant not assigned: no lineage assigned due to poor - sequence quality **OR** low viral load (ct≥35) **OR** variant PCR could not assign variant and no sequencing result

The majority of ILI cases were of black race (71.9%, 1972/2744) and 54.7% (1501/2742) were female. Eight percent (207/2670) of ILI cases with known COVID-19 vaccination status were fully vaccinated at the time of enrolment. On multivariable analysis among individuals with ILI, SARS-CoV-2-positive cases were more likely to be in older age groups (45-64 years [adjusted odds ratio (aOR) 1.4, 95% confidence interval (CI) 1.1-1.9] and ≥65 years [aOR 1.9, 95% CI 1.1-3.1]) compared with 15-24 years, to present during the wave (aOR 3.4, 95%CI 2.7-4.2) compared to out of wave and to present ≥5 (aOR 1.3, 95% CI 1.1-1.5) days from date of symptom onset compared to <5 days. SARS-CoV-2 cases were less likely to be co-infected with RSV (aOR 0.2, 95% CI 0.1-0.6) and to have asthma (aOR 0.4, 95%CI 0.2-0.8).

The majority of SRI cases were of black race (81.3%, 5928/7014) and 55.7% (3905/7013) were female. Five percent (342/6884) of cases with known COVID-19 vaccination status were fully vaccinated. On multivariable analysis, among individuals with SRI (including all 8 surveillance hospitals), SARS-CoV-2-positive cases were more likely to be 45-64 years (aOR 1.5, 95% CI 1.1-2.1) compared with 15-24 years, to be diabetic (aOR 1.4, 95%CI 1.2-1.7) or obese (aOR 1.4, 95% CI 1.2-1.7), to be enrolled during a wave compared to out of wave (aOR 4.6, 95%CI 4.0-5.3), to receive oxygen therapy (aOR 1.4, 95%CI 1.2-1.6), be admitted to an intensive care unit (ICU) (aOR 6.8, 95%CI 3.5-13.2) and to die (aOR 1.5, 95% CI 1.2-1.7) during admission. SARS-CoV-2 cases were less likely to be male (aOR 0.7, 95%CI 0.6-0.8), to be infected with HIV (aOR 0.6, 95%CI 0.5-0.7), to have asthma (aOR 0.6, 95%CI 0.4-0.7) or have previous or current tuberculosis (aOR 0.2, 95%CI 0.1-0.3) compared to those testing SARS-CoV-2 negative.

### Risk factors for SARS-CoV-2 associated hospitalisation

From 01 April 2020 through 31 March 2022, a total of 2746 ILI and 3905 SRI cases were enrolled at the three sentinel sites (Western Cape, North West and KwaZulu-Natal) with surveillance for SRI and ILI in the same catchment population. SARS-CoV-2 was detected in 24.0% (660/2746) and 36.9% (1439/3905) of ILI and SRI cases, respectively. HIV prevalence was 19.4% (125/643) and 25.0% (345/1378) among SARS-CoV-2 positive-ILI and -SRI cases, respectively (Table 2).

**Table 2:**
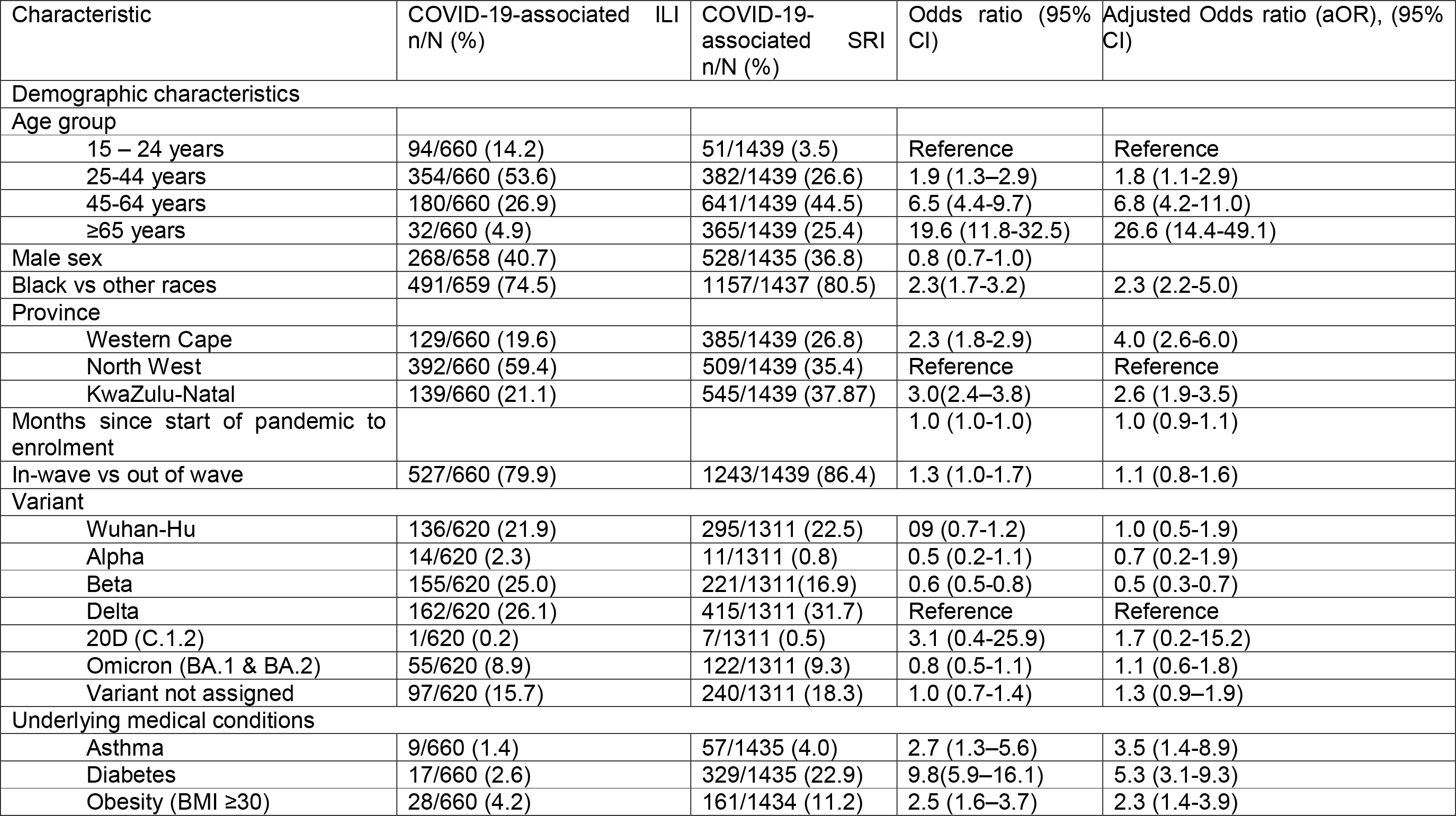
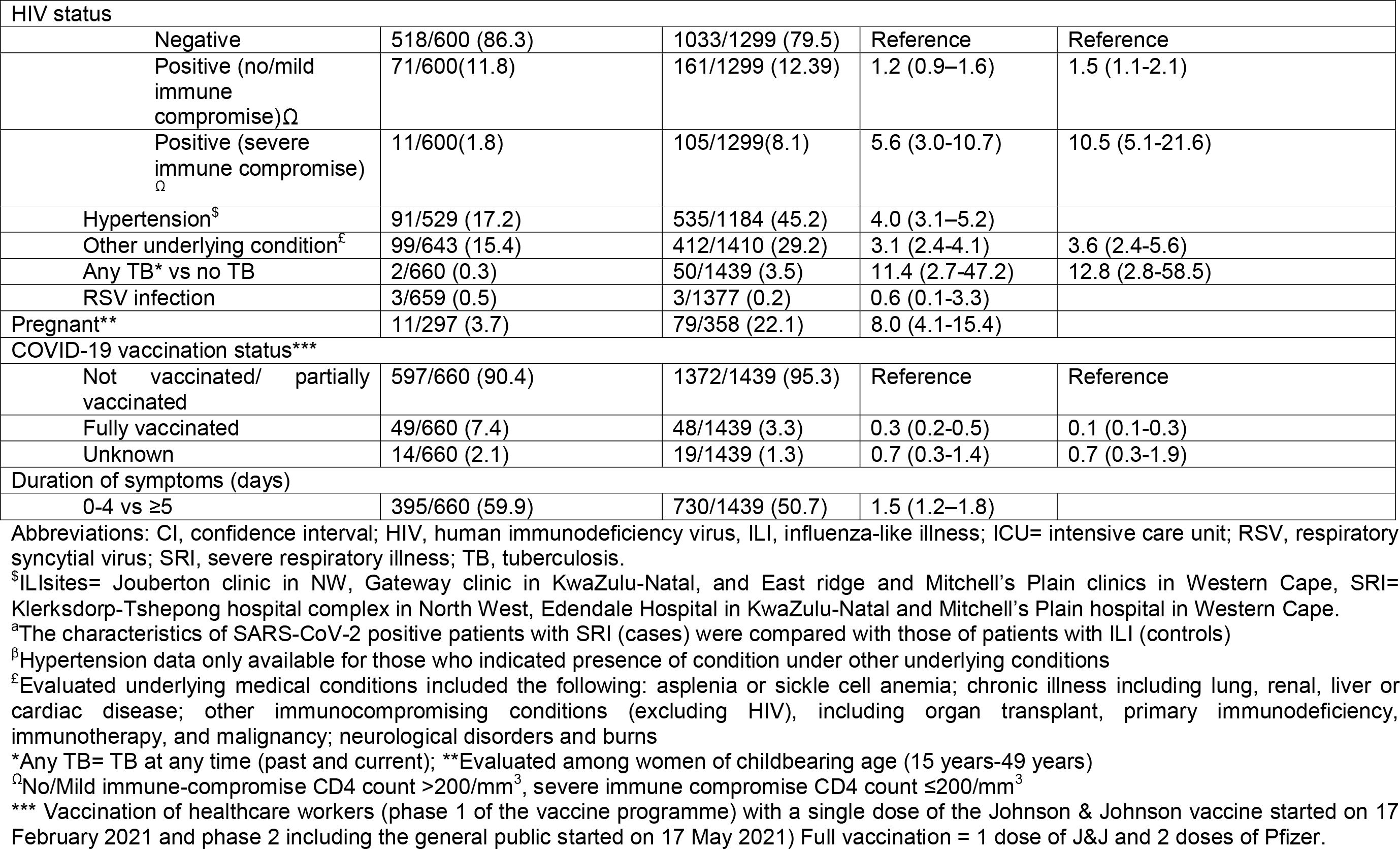
Risk factors for COVID-19-associated hospitalisation among persons aged ≥15 years at 3 sentinel sites^$^ in South Africa, April 2020 – March 2022 N=6651.

| Characteristic | COVID-19-associated ILI<br>n/N (%) | COVID-19-associated SRI<br>n/N (%) | Odds ratio (95%<br>CI) | Adjusted Odds ratio (aOR), (95%<br>CI) |
| --- | --- | --- | --- | --- |
| **Demographic characteristics** |  |  |  |  |
| Age group |  |  |  |  |
| 15 – 24 years | 94/660 (14.2) | 51/1439 (3.5) | Reference | Reference |
| 25-44 years | 354/660 (53.6) | 382/1439 (26.6) | 1.9 (1.3–2.9) | 1.8 (1.1-2.9) |
| 45-64 years | 180/660 (26.9) | 641/1439 (44.5) | 6.5 (4.4-9.7) | 6.8 (4.2-11.0) |
| ≥65 years | 32/660 (4.9) | 365/1439 (25.4) | 19.6 (11.8-32.5) | 26.6 (14.4-49.1) |
| Male sex | 268/658 (40.7) | 528/1435 (36.8) | 0.8 (0.7-1.0) |  |
| Black vs other races | 491/659 (74.5) | 1157/1437 (80.5) | 2.3(1.7-3.2) | 2.3 (2.2-5.0) |
| Province |  |  |  |  |
| Western Cape | 129/660 (19.6) | 385/1439 (26.8) | 2.3 (1.8-2.9) | 4.0 (2.6-6.0) |
| North West | 392/660 (59.4) | 509/1439 (35.4) | Reference | Reference |
| KwaZulu-Natal | 139/660 (21.1) | 545/1439 (37.87) | 3.0(2.4–3.8) | 2.6 (1.9-3.5) |
| Months since start of pandemic to enrolment |  |  | 1.0 (1.0-1.0) | 1.0 (0.9-1.1) |
| In-wave vs out of wave | 527/660 (79.9) | 1243/1439 (86.4) | 1.3 (1.0-1.7) | 1.1 (0.8-1.6) |
| **Variant** |  |  |  |  |
| Wuhan-Hu | 136/620 (21.9) | 295/1311 (22.5) | 09 (0.7-1.2) | 1.0 (0.5-1.9) |
| Alpha | 14/620 (2.3) | 11/1311 (0.8) | 0.5 (0.2-1.1) | 0.7 (0.2-1.9) |
| Beta | 155/620 (25.0) | 221/1311(16.9) | 0.6 (0.5-0.8) | 0.5 (0.3-0.7) |
| Delta | 162/620 (26.1) | 415/1311 (31.7) | Reference | Reference |
| 20D (C.1.2) | 1/620 (0.2) | 7/1311 (0.5) | 3.1 (0.4-25.9) | 1.7 (0.2-15.2) |
| Omicron (BA.1 & BA.2) | 55/620 (8.9) | 122/1311 (9.3) | 0.8 (0.5-1.1) | 1.1 (0.6-1.8) |
| Variant not assigned | 97/620 (15.7) | 240/1311 (18.3) | 1.0 (0.7-1.4) | 1.3 (0.9–1.9) |
| **Underlying medical conditions** |  |  |  |  |
| Asthma | 9/660 (1.4) | 57/1435 (4.0) | 2.7 (1.3–5.6) | 3.5 (1.4-8.9) |
| Diabetes | 17/660 (2.6) | 329/1435 (22.9) | 9.8(5.9–16.1) | 5.3 (3.1-9.3) |
| Obesity (BMI ≥30) | 28/660 (4.2) | 161/1434 (11.2) | 2.5 (1.6–3.7) | 2.3 (1.4-3.9) |

|  |  |  |  |  |  |
| --- | --- | --- | --- | --- | --- |
| HIV status |  |  |  |  |  |
| Negative | 518/600 (86.3) | 1033/1299 (79.5) | Reference | Reference |  |
| Positive (no/mild immune compromise)<sup>Ω</sup> | 71/600(11.8) | 161/1299 (12.39) | 1.2 (0.9–1.6) | 1.5 (1.1-2.1) |  |
| Positive (severe immune compromise)<sup>Ω</sup> | 11/600(1.8) | 105/1299(8.1) | 5.6 (3.0-10.7) | 10.5 (5.1-21.6) |  |
| Hypertension<sup>\$</sup> | 91/529 (17.2) | 535/1184 (45.2) | 4.0 (3.1–5.2) | | |
| Other underlying condition<sup>£</sup> | 99/643 (15.4) | 412/1410 (29.2) | 3.1 (2.4-4.1) | 3.6 (2.4-5.6) |  |
| Any TB* vs no TB | 2/660 (0.3) | 50/1439 (3.5) | 11.4 (2.7-47.2) | 12.8 (2.8-58.5) |  |
| RSV infection | 3/659 (0.5) | 3/1377 (0.2) | 0.6 (0.1-3.3) |  |  |
| Pregnant** | 11/297 (3.7) | 79/358 (22.1) | 8.0 (4.1-15.4) |  |  |
| COVID-19 vaccination status*** |  |  |  |  |  |
| Not vaccinated/ partially vaccinated | 597/660 (90.4) | 1372/1439 (95.3) | Reference | Reference |  |
| Fully vaccinated | 49/660 (7.4) | 48/1439 (3.3) | 0.3 (0.2-0.5) | 0.1 (0.1-0.3) |  |
| Unknown | 14/660 (2.1) | 19/1439 (1.3) | 0.7 (0.3-1.4) | 0.7 (0.3-1.9) |  |
| Duration of symptoms (days) |  |  |  |  |  |
| 0-4 vs ≥5 | 395/660 (59.9) | 730/1439 (50.7) | 1.5 (1.2–1.8) |  |  |

Abbreviations: CI, confidence interval; HIV, human immunodeficiency virus; ILI, influenza-like illness; ICU= intensive care unit; RSV, respiratory syncytial virus; SRI, severe respiratory illness; TB, tuberculosis.
<sup>\$</sup>ILIsites= Jouberton clinic in NW, Gateway clinic in KwaZulu-Natal, and East ridge and Mitchell's Plain clinics in Western Cape, SRI= Klerksdorp-Tshepong hospital complex in North West, Edendale Hospital in KwaZulu-Natal and Mitchell's Plain hospital in Western Cape.
<sup>a</sup>The characteristics of SARS-CoV-2 positive patients with SRI (cases) were compared with those of patients with ILI (controls)
<sup>β</sup>Hypertension data only available for those who indicated presence of condition under other underlying conditions
<sup>£</sup>Evaluated underlying medical conditions included the following: asplenia or sickle cell anemia; chronic illness including lung, renal, liver or cardiac disease; other immunocompromising conditions (excluding HIV), including organ transplant, primary immunodeficiency, immunotherapy, and malignancy; neurological disorders and burns
\*Any TB= TB at any time (past and current); \*\*Evaluated among women of childbearing age (15 years-49 years)
<sup>Ω</sup>No/Mild immune-compromise CD4 count >200/mm<sup>3</sup>, severe immune compromise CD4 count ≤200/mm<sup>3</sup>
\*\*\* Vaccination of healthcare workers (phase 1 of the vaccine programme) with a single dose of the Johnson & Johnson vaccine started on 17 February 2021 and phase 2 including the general public started on 17 May 2021) Full vaccination = 1 dose of J&J and 2 doses of Pfizer.

On multivariable analysis, comparing SARS-CoV-2-positive SRI cases to SARS-CoV-2-positive ILI cases, factors associated with increased risk of COVID-19-associated hospitalisation included older age (25-44 years [aOR 1.8, 95%CI 1.1-2.9], 45-64 years [aOR 6.8, 95%CI 4.2-11.0] and ≥65 years [aOR 26.6, 95%CI 14.4-49.1] compared to 15-24 years), being of black race (aOR 3.3, 95% CI 2.2-5.0), obesity (aOR 2.3, 95% CI 1.4-3.9), asthma (aOR 3.5, 95% CI 1.4-8.8), diabetes (aOR 5.3, 95%CI 3.1-9.3), previous or current tuberculosis (aOR 12.8, 95%CI 2.9-58.5) or HIV with CD4 ≥200/mm^3^ (aOR 1.5, 95%CI 1.1-2.2) and CD4<200/mm^3^ (aOR 10.5, 95%CI 5.1-21.6); being enrolled from KwaZulu-Natal (aOR 2.6, 95%CI 1.9-3.6) or Western Cape (aOR 4.0, 95% CI 2.6-6.0) compared to North West Province. Infection with Beta variant (aOR 0.5, 95% CI 0.3-0.7) compared to Delta variant and being fully vaccinated vs not vaccinated/partially vaccinated (aOR 0.1, 95%CI 0.1-0.3) were less likely to be associated with COVID-19-associated SRI hospitalisation.

### Risk factors for COVID-19-associated mortality

During the study period, 7025 patients with SRI were enrolled at the surveillance hospital sites, of which 2282 (32.5%) tested positive for SARS-CoV-2. Of the 2255 COVID-19 patients with in-hospital outcome data, 387 (17.2%) died (Table 3). The median age of SARS-CoV-2 patients who died was 62.3 years (interquartile range (IQR) 52.1-69.5 years), 55.8% (216/387) were female and 24.5% (87/355) were PLHIV. Older age (45-64 years [aOR 2.6, 95% CI 1.7-3.9] and ≥65 years [aOR 4.7, 95% CI 3.1-7.3] compared to 25-44 years), admission for <3 days (aOR 1.6, 95% CI 1.1-2.2) compared to 3-7 days were associated with in-hospital mortality. Infection with Omicron (BA.1 and BA.2) [aOR 0.3, 95%CI, 0.2-0.6] compared to Delta variant was less likely to be associated with in-hospital mortality.

**Table 3:** Factors associated with mortality among patients aged ≥15 years hospitalised with severe respiratory illness (SRI) testing positive for SARS-CoV-2, at 5 sentinel sites in South Africa, April 2020-March 2022, n=2148.

| Characteristic | Case fatality ratio n/N (%) | Odds Ratio (95% CI) | Adjusted Odds ratio (aOR), (95% CI) |
| --- | --- | --- | --- |
| Age groups |  |  |  |
| 15-24 years | 2/75 (2.7) | 0.3 (0.1-1.2) | 0.3 (0.9-1.6) |
| 25-44 years | 53/635 (8.4) | Reference | Reference |
| 45-64 years | 169/968 (17.5) | 2.3 (1.7-3.2) | 2.2 (1.6-3.2) |
| ≥65 years | 163/577 (28.3) | 4.1 (2.9-5.7) | 4.0 (2.8-5.8) |
| Sex |  |  |  |
| Female | 216/1389 (15.6) | Reference | Reference |
| Male | 171/861 (19.9) | 1.4 (1.1-1.7) | 1.3 (1.0-1.6) |
| Race |  |  |  |
| Other | 66/413 (15.6) | Reference |  |
| Black | 321/1839 (17.5) | 0.8 (0.6-1.2) |  |
| Province |  |  |  |
| North West | 62/509 (12.8) | 0.8 (0.5-1.2) | 0.9 (0.4-1.7) |
| Mpumalanga | 68/275 (24.7) | 1.9 (1.3-2.9) | 2.0 (1.4-3.1) |
| KwaZulu-Natal | 126/544 (23.2) | 1.8 (1.2-2.5) | 1.8 (1.2-2.8) |
| Western Cape | 54/370 (14.6) | Reference | Reference |
| Gauteng | 77/557 (13.8) | 0.9 (0.6 – 1.4) | 1.3 (0.9-2.0) |
| Wave vs out of wave |  |  | 1.0 (0.7-8.5) |
| Period since 1<sup>st</sup> case in South Africa (months) |  | 1.0 (1.0-1.0) | 1.0 (0.9-1.1) |
| Variant |  |  |  |
| Wuhan-Hu | 66/445 (14.8) | 0.6 (0.4-0.9) | 0.8 (0.4-1.7) |
| Alpha | 4/19 (21.1) | 1.1 (0.3-3.3) | 1.2 (0.4-3.8) |
| Beta | 95/437 (21.7) | 0.9 (0.7-1.2) | 1.0 (0.6-1.5) |
| Delta | 133/577 (23.1) | Reference | Reference |
| Omicron (BA.1 & BA.2) | 17/197 (8.6) | 0.3 (0.2-0.5) | 0.3 (0.2-0.6) |
| 20D (C.1.2) | 5/14 (35.7) | 2.0 (0.7-6.3) | 1.9 (0.6-6.2) |
| Variant not assigned | 40/380(10.5) | 0.4 (0.3-0.6) | 0.5 (0.3-0.7) |
| Asthma |  |  |  |
| No | 360/2066 (17.4) | Reference |  |
| Yes | 12/76 (15.8) | 0.9 (0.5-1.6) |  |
| Diabetes |  |  |  |
| No | 288/1777 (16.2) | Reference |  |
| Yes | 99/472 (21.0) | 1.3 (1.0 – 1.7) |  |
| Obesity (BMI ≥30) |  |  |  |
| No | 335/ 2010 (16.7) | Reference |  |
| Yes | 52/ 238 (21.9) | 1.3 (0.9 – 1.8) |  |
| HIV status |  |  |  |
| Negative | 268/1511 (17.6) | Reference |  |
| Positive (no/mild immune suppression)<sup>Ω</sup> | 27/218 (12.4) | 0.6 (0.4-1.0) |  |
| Positive (severe immune suppression)<sup>Ω</sup> | 23/170 (13.5) | 0.8 (0.5-1.3) |  |
| Hypertension |  |  |  |
| No | 164/1055 (15.6) | Reference |  |
| Yes | 162/757 (21.4) | 1.5 (1.2-1.9) |  |
| Tuberculosis |  |  |  |
| No TB | 379/2174 (17.4) | Reference |  |
| Any TB* | 8/81 (9.9) | 0.5 (0.2-1.0) |  |
| RSV infection |  |  |  |
| No | 370 /2177 (17.0) | Reference |  |
| Yes | 2/9 (22.2) | 1.3 (0.3-6.5) |  |
| Other underlying condition<sup>‡</sup> |  |  |  |
| No | 318/1833 (17.4) | Reference |  |
| Yes | 69/422 (16.4) | 1.0 (0.8-1.3) |  |
| Pregnant |  |  |  |
| No | 215/1223 (17.6) | Reference |  |
| Yes | 1/164 (0.6) | 0.03 (0.00-0.2) |  |
| COVID-19 vaccination status** |  |  |  |
| Not vaccinated/ partially vaccinated | 373/2160(17.2) | Reference | Reference |
| Fully vaccinated | 8/69 (11.6) | 0.6 (0.3-1.2) | 0.8 (0.3-1.8) |
| Unknown | 6/26 (23.1) | 1.2 (0.6-4.0) | 1.3 (0.5-3.9) |
| Duration of symptoms (days) |  |  |  |
| 0-4 days | 174/1074 (15.8) | Reference |  |
| ≥5 | 213/1157 (18.4) | 1.1 (0.9–1.4) |  |
| Duration of hospitalisation (days) |  |  |  |
| <3 | 250/1357 (18.4) | 1.6 (1.2-2.1) | 1.7 (1.2-2.2) |
| 3-7 | 358/3034 (11.8) | Reference | Reference |
| ≥8 | 331/ 2522 (13.1) | 1.0 (0.7-1.2) | 1.0 (0.7-1.3) |

Abbreviations: CI, confidence interval; HIV, human immunodeficiency virus; ILI, influenza-like illness; ICU= intensive care unit; RSV, respiratory syncytial virus; SRI, severe respiratory illness; TB, tuberculosis.
<sup>β</sup>Hypertension was not collected as a standalone variable; patients reported it under other underlying conditions
<sup>ε</sup>Evaluated underlying medical conditions included the following: asplenia or sickle cell anemia; chronic illness including lung, renal, liver or cardiac disease; other immunocompromising conditions (excluding HIV), including organ transplant, primary immunodeficiency, immunotherapy, and malignancy; neurological disorders and burns
\*Any TB= TB at any time (past and current)
<sup>Ω</sup>No/mild immune suppression= CD4 count >200/mm<sup>3</sup>, severe immune suppression =CD4 count ≤200/mm<sup>3</sup>
\*\* Vaccination of healthcare workers (phase 1 of the vaccine programme) with a single dose of the Johnson & Johnson vaccine started on 17 February 2021 and phase 2 including the general public started on 17 May 2021) Full vaccination = 1 dose of J&J and 2 doses of Pfizer.

## Discussion

Using data from enhanced routine sentinel surveillance, we provide data on risk factors for COVID-19–associated hospitalisation and mortality among individuals aged 15 years and older in a high HIV and high tuberculosis prevalence setting. Older age was strongly associated with hospitalisation and mortality. In addition to older age, prior or current tuberculosis and severe HIV immunosuppression were associated with increased risk of COVID-19 associated hospitalisation. Our study showed that compared to the Delta variant, the Omicron variant (BA.1 and BA.2) was associated with reduced risk of mortality and Beta associated with decreased risk of hospitalisation. Similar to other studies, older age and underlying conditions (diabetes, obesity, asthma, HIV) were associated with severe COVID-19 illness[17, 18].

Compared to other causes of respiratory illness, SARS-CoV-2 was more likely to affect older individuals and was less likely to affect PLHIV. HIV infection has been associated with increased risk of severe illness associated with influenza[19] and bacterial pathogens such as the pneumococcus[20]. The apparent negative association with SARS-CoV-2 detection amongst SRI cases is likely because non- or mildly immunocompromised HIV may be less of a risk factor for SRI due to SARS-CoV-2 compared to other respiratory pathogens in the comparison group.

In this study, we identified risk factors for COVID-19 -associated hospitalisation which were similar to what has been described by other studies, such as older age, asthma, diabetes mellitus, tuberculosis and obesity [5, 17, 18, 21, 22]. In addition, increasing level of HIV immune compromise was also associated with higher risk of COVID-19 hospitalisation. Population studies conducted during the first wave of SARS-CoV-2 infections, reported an increased risk of hospitalisation or mortality among PLHIV as compared to the general population [18, 23]. A population cohort study covering the first wave of infections in the Western Cape Province of South Africa, reported increased risk of mortality (adjusted hazard ratio [aHR], 2.14) among PLHIV compared to the general population and that CD4 <200 cells/μL during current admission was associated with mortality [18]. Similarly, a population-based study in England, reported that HIV-infection was associated with higher risk of death due to COVID-19 (aHR 2·90, 95%CI 1·96–4·30). In a cohort study in South Africa, among cases hospitalised with COVID-19, PLHIV with a history of immune compromise (CD4 count <200 cells/μL) were more likely to die in-hospital than those with CD4 counts of ≥200 cells/μL. However, CD4 count was only available for 20% of individuals[24]. Similarly, a retrospective cohort study using routinely collected data showed that after adjusting for sex and age, SARS-CoV-2 positive PLHIV compared to HIV-uninfected had higher risk of death (hazard ratio 2.9, 95%CI 2.0-4.2)[23].

Our study used a novel approach, leveraging long-standing syndromic sentinel surveillance for influenza and systematically collected epidemiological and SARS-CoV-2 genomic data and comparing data from outpatients and inpatients. This enhanced syndromic sentinel surveillance reported similar trends to those reported by the two national surveillance systems used in South Africa in response to COVID-19, the national laboratory-based surveillance and Data for COVID-19 (DATCOV) national active surveillance for COVID-19 hospitalization[2, 24]. The common trends included the timing of the four waves of infection, each dominated by a different variant for the period under surveillance, higher prevalence of SARS-CoV-2 infection among females, and individuals of black race as compared to other races and similar risk factors for severe illness including older age and underlying illness such as tuberculosis and HIV with severe immunocompromise. In addition, the dominant variants in the waves also corresponded to the national data. Because sentinel surveillance provided generally robust data for tracking the SARS-CoV-2 pandemic comparable to data from national surveillance, it may represent a viable surveillance approach in settings where national surveillance is not possible and may also be a sustainable option for moving beyond the pandemic phase when testing for SARS-CoV-2 is reduced. The World Health Organization is encouraging countries to use systematically collected surveillance data to monitor the SARS-CoV-2 pandemic, and the guidance on the end- to- end integration of influenza and COVID-19 sentinel surveillance has been published[25].

In our sentinel surveillance, 17.2% of hospitalised cases with laboratory-confirmed COVID-19 died, and in-hospital mortality was highest among COVID-19 patients aged 65 years and older. This was slightly lower than the 26.2% reported for public sector hospitals included in the DATCOV national surveillance[26], most likely due to the fact that our surveillance system required patients to sign consent for participation and critically ill patients may therefore have been missed. For the mortality model, we were not able to show a significant association with some risk factors reported by other studies, such as tuberculosis, or some underlying conditions (obesity, asthma, hypertension, chronic cardiac and pulmonary conditions) possibly because numbers enrolled were too small to detect an association. This lack of power, especially for mortality analysis, represents a potential weakness of the sentinel surveillance approach that requires individual patient consent when compared to national surveillance. However, pooled analysis across different countries may be valuable.

Our study showed that the Omicron variant, which emerged in November 2021 in South Africa[27, 28], was associated with lower risk of mortality compared to the Delta variant. Our findings are similar to other studies [29-32]. Results from an analysis using data from national case, hospitalisations and genomic surveillance systems, showed that compared with Delta variant infections, S gene target failure-infections (proxy for Omicron variant) had a significantly lower odds of severe disease (aOR 0·3, 95% CI 0·2–0·5) among hospitalised individuals in South Africa [29].

The strengths of our study include that data were collected systematically in both inpatient and outpatient populations using an existing surveillance platform and we were able to compare cases presenting with mild disease to those who were admitted. In addition, data on SARS-CoV-2 lineage was available for a majority of positive cases.

Our study has some limitations. Our surveillance covers five of the nine provinces, including three of the most populous provinces, in South Africa, and data from this study may not be generalizable to the rest of the country, especially if transmission patterns vary across provinces. For example, the phylogeographic analysis suggests that Beta variant which was the driver of the second wave in South Africa emerged in early August in Eastern Cape Province (which is not covered by surveillance) [33] but this variant was not detected in our enhanced sentinel surveillance until the first week of November 2020. Our study only collected data on in-hospital mortality, it is possible that patients could have died after discharge from hospital or following ILI consultation.

We demonstrate that an existing sentinel surveillance system could be modified to monitor the COVID-19 pandemic and likely any future pandemics once community transmission has been established. We described risk factors associated with SARS-CoV-2-associated hospitalisation and mortality in a setting of high HIV and tuberculosis prevalence. Active syndromic surveillance combining clinical, laboratory and genomic data can be used to describe the epidemic timing, epidemiological characteristics of cases, early detection of variants of concern and how these impact disease severity and outcomes. The association of severe HIV-associated immune compromise with increased hospitalisation highlights the importance of increasing access to antiretroviral therapy and this group should be prioritised for vaccination. In addition, the increased risk of hospitalisation and mortality among individuals aged ≥65 years supports South Africa’s Department of Health guidance of including individuals aged >60 years as a the priority group for COVID-19 vaccination and booster doses [34].

## Data Availability

All data produced in the present study are available upon reasonable request to the authors.

## Acknowledgments

We would like to thank the participants in the surveillance program for their time and patience, as well as the ILI, pneumonia, GERMS-SA surveillance officers and research assistants; Centre for Respiratory Diseases and Meningitis laboratory team and data team. We also acknowledge the following site investigators:

Dr Omphile Mekgoe, Department of Paediatrics, Klerksdorp Hospital, Klerksdorp, South Africa

Dr Godfrey Siwele, Department of Medicine, Matikwana Hospital, Hazy View, South Africa

Dr Neydis Baute, Department of Paediatrics, Mapulaneng hospital, Hazy View, South Africa

## Disclaimer

The findings and conclusions in this report are those of the authors and do not necessarily represent the official position of the US Centers for Disease Control and Prevention.

