## supplemental material for "Risk factors for severe COVID-19 among HIV-infected and-uninfected individuals in South Africa, April 2020- March 2022 – data from sentinel surveillance"

**Methods**

**Laboratory Procedures**

From 1 April 2020 – 28 February 2021, samples were tested for SARS-CoV-2 using the TIB MOLBIOL E gene assay (Roche Diagnostics)[1]. Influenza and RSV were detected using the Fast Track Diagnostics (FTD) Flu/HRSV kit (Siemens). From 1 March 2021, samples were tested using the Allplex™ SARS-CoV-2/Flu A/Flu B/RSV kit (Seegene, Seoul, South Korea). SARS-CoV-2 positivity was assigned if the PCR cycle threshold (C_t_) value was <40 for ≥1 target (N, S or RdRp). Variants of concern (VOC) were determined by PCR as follows: from 1 April 2020 to 30 June 2021, SARS-CoV-2 samples were typed using the Allplex™ Variants I typing assay which detects Alpha and Beta/Gamma VOCs. In addition, samples collected from 1 January – 30 June 2021 were run on the Allplex™ Variants II assay which detects the Delta variant and differentiates Beta from Gamma. In addition to PCR, SARS-CoV-2 positive samples were sequenced to ascertain their lineage/clade. From July 2021, onwards, only sequencing was used to ascertain their lineage/clade.

For sequencing, briefly, RNA was extracted either manually or automatically in batches, using the QIAamp viral RNA mini kit (QIAGEN, CA, USA) or the Chemagic 360 using the CMG-1049 kit (PerkinElmer, MA, USA). Sequencing was performed with the amplicon-based next-generation sequencing approaches using the Illumina COVIDSeq protocol (Illumina Inc., CA, USA) or nCoV-2019 ARTIC network sequencing protocol v3 (https://artic.network/ncov-2019). Raw reads from Illumina sequencing were assembled using the Exatype NGS SARS-CoV-2 pipeline v1.6.1, (https://sars-cov-2.exatype.com/). The resulting consensus sequence was further manually polished by considering and correcting indels in homopolymer regions that break the open reading frame (probably sequencing errors) using Aliview v1.27, (http://ormbunkar.se/aliview/)[2]. All assemblies determined to have acceptable quality (defined as having at least 1 000 000 reads and at least 50 % 10 X coverage) were deposited on GISAID (https://www.gisaid.org/)[3, 4]). Assembled genomes were assigned lineages using the ‘Phylogenetic Assignment of Named Global Outbreak Lineages’ (PANGOLIN) software suite (https://github.com/hCoV-2019/pangolin)[5]. The SARS-CoV-2 genomes were also classified using the clade classification proposed by NextStrain (https://nextstrain.org/)[6].

**Ethical approval**

In addition to the University of Witwatersrand Human Research Ethics Committee (HREC) approval, the ILI protocol was approved by the University of KwaZulu-Natal Human Biomedical Research Ethics Committee (BREC) reference BF 080/12 and the University of Cape Town Faculty of Health Science Human Research Ethics Committee (FHS HREC) reference 573/2018. In addition to the approval by the University of Witwatersrand HREC, the SRI protocol was approved by University of Cape Town FHS HREC reference 836/2014, and BREC reference M496/14.
